# Predictors of Mortality in Hospitalized Patients with COVID-19: A One-Year Case-Control Study

**DOI:** 10.1101/2023.05.12.23289918

**Authors:** Laura Camacho-Domínguez, Manuel Rojas, María Herrán, Yhojan Rodríguez, Santiago Beltrán, Paola Saboya Galindo, Nicolas Aguirre-Correal, María Espitia, Santiago García, Valeria Bejarano, Victoria Morales-González, Jaime Enrique Covaleda-Vargas, Mónica Rodríguez-Jiménez, Elizabeth Zapata, Diana M. Monsalve, Yeny Acosta-Ampudia, Juan-Manuel Anaya, Carolina Ramírez-Santana

## Abstract

**Objective:** To determine the associated factors with mortality, in addition to age and sex, in a high-complexity hospital in Bogota, Colombia, during the first year of the pandemic.

**Design:** A case-control study.

**Setting:** High-complexity center above 2,640 meters above sea level (masl) in Colombia.

**Methods:** A case-control study was conducted on 564 patients admitted to the hospital with confirmed COVID-19. Deceased patients (n: 282) and a control group (n: 282), matched by age, sex, and month of admission, were included. Clinical and paraclinical variables were retrospectively obtained by systematic revision of clinical records. Multiple imputations by chained equation (MICE) were implemented to account for missing variables. Classification and regression trees (CART) were estimated to evaluate the interaction of associated factors on admission and their role in predicting mortality during hospitalization.

**Results:** Most of the patients included were males in the seventh decade of life. Most of the admissions occurred between July and August 2021. Surprisingly, recovered patients reported heterogeneous symptomatology, whereas deceased patients were most likely to present respiratory distress, dyspnea, and seizures on admission. In addition, the latter group exhibited a higher burden of comorbidities and alterations in laboratory parameters. After the imputation of datasets, CART analysis estimated 14 clinical profiles based on respiratory distress, LDH, dyspnea, hemoglobin, D-dimer, ferritin, blood urea nitrogen, C-reactive protein, PaO_2_/FiO_2_, dysgeusia, total bilirubin, platelets, and gastroesophageal reflux disease. The accuracy model for prediction was 85.6% (P < 0.0001).

**Conclusion:** Multivariate analysis yielded a reliable model to predict mortality in COVID-19. This analysis revealed new interactions between clinical and paraclinical features in addition to age and sex. Furthermore, this predictive model could offer new clues for the personalized management of this condition in clinical settings.

## Introduction

Coronavirus disease 2019 (COVID-19), caused by the severe acute respiratory syndrome coronavirus 2 (SARS-CoV-2), was first reported as an outbreak of new viral pneumonia in Wuhan, China which was quickly distributed worldwide, generating a remarkable impact. Approximately 520,000,000 cases have been reported, from which about 6,300,000 have died (1). This infection has a wide range of clinical manifestations, ranging from asymptomatic disease to individuals who develop acute respiratory failure (2). However, most patients have mild symptoms of cough, headache, myalgia, fever, and diarrhea; a smaller proportion presents severe disease symptoms (1,2). The most common manifestation of severity is dyspnea, which can be seen in up to 40% of patients and is usually accompanied by hypoxemia (2).

The United States harbors almost a fifth of the infections worldwide and more than 1,000,000 deaths (1). It is followed by Brazil and India, with 665,000 and 524,000 deaths, respectively (1). Colombia occupies the twelfth position with approximately 140,000 deaths (1,3). Fatality, defined as the ratio between the cases of mortality and the cases confirmed with SARS-CoV-2, has progressively decreased from 3.7% in March 2020 to 1.22% in the present day (4). Colombia presents a fatality rate of 2.5%, higher than reported worldwide (3). Thus, further characterization of mortality in Colombia is required to develop personalized risk profiles to better respond to the pandemic, especially in the current transition to endemic disease.

Patients with severe diseases are more likely to suffer complications associated with higher mortality. One of the causes of this clinical deterioration is the cytokine storm that generates a systemic inflammatory syndrome in which high cytokine levels are associated with hyperactivation of the cellular response (5–8). Molecular mimicry between host and viral proteins has also been described, as well as direct damage of the microorganism to tissues that present high expression of the angiotensin-converting enzyme-2 receptor, as other possible causes (5–8). These processes will result, in most cases, in a respiratory distress syndrome characterized by impaired gas exchange (6,8). They could also generate a hypercoagulability state responsible for the increase in thrombotic events in patients with this condition (5,7,9).

Multiple studies have found several factors associated with developing severe disease. Age, sex, ethnicity, socioeconomic status, inflammatory markers, and comorbid conditions have been widely associated with mortality. However, other factors include ancestry, environmental exposures, viral mutations, and geographic diversity (10–12). A recent study in Bogota, a city above 2,640 meters above sea level (masl) in Colombia, found that older age, low S/F: Ratio of oxygen saturation to fraction-inspired of oxygen (S/F), and high lactate dehydrogenase (LDH) on admission were predictors of mortality (13). However, observational studies have shown that other variables, in addition to age and sex, could exhibit interactions among them, which have been poorly characterized in our population (14).

Herein, we conducted a one-year case-control study to characterize the clinical and paraclinical factors associated with mortality during the first year of the pandemic. We implemented supervised machine learning algorithms to evaluate the interactions among variables and their potential to predict mortality on admission. We adjusted the analysis by monthly progression, which may account for changes in the mortality rates due to viral mutations during the pandemic.

## Methods

### Study design

This case-control study was conducted in Clínica de Occidente, a tertiary referral center in Bogota, Colombia. The study screening was from February 29^th^, 2020, to March 1^st^, 2021. Included patients were selected from 4,163 cases positive for SARS-CoV-2 by reverse transcriptase-polymerase chain reaction (RT-PCR) and who attended the emergency room during the first year of the pandemic with complete clinical records. A non-probabilistic convenience sampling included a total of 282 deceased patients. An additional group of 282 patients, who were hospitalized but recovered from COVID-19, was included as the control group. These controls were matched to deceased patients by age, sex, and month of consultation (to account for viral mutations). None of the patients received vaccinations during the study timeframe, and the included patients attended the emergency room between April 3^rd^, 2020, to January 30^th^, 2021. This was a low-risk study according to the resolution 8430 of 1993 from the Ministry of Health of Colombia. The institutional review board of Clínica de Occidente approved the study design.

### Inclusion and exclusion criteria

Inclusion criteria comprised the following: (1) aged at least 18 years; (2) COVID-19 diagnosis based on RT-PCR testing; (3) hospitalized patients; and (4) death registered in death certificate adequately filled out by medical doctors who certified COVID-19 as the cause of death. In the case of controls, they were hospitalized but recovered during the hospitalization and received ambulatory management. Patients referred to another clinical setting were excluded from the analysis.

### Clinical variables and data extraction

Medical records were reviewed through access to the server assigned by the hospital. Information considered relevant for this investigation was extracted, including sociodemographic variables, clinical features (signs, symptoms, laboratories on admission), past medical history, in-hospital management (i.e., requirement of ventilatory support, intensive care unit (ICU) admission, vasopressor support), medications administered on admission, organ-specific and systemic outcomes during hospitalization (Table 1, 2 and 3). Research participants remained anonymous at all times. All data were collected in an electronic and secure database as described elsewhere (15).

**Table 1.**
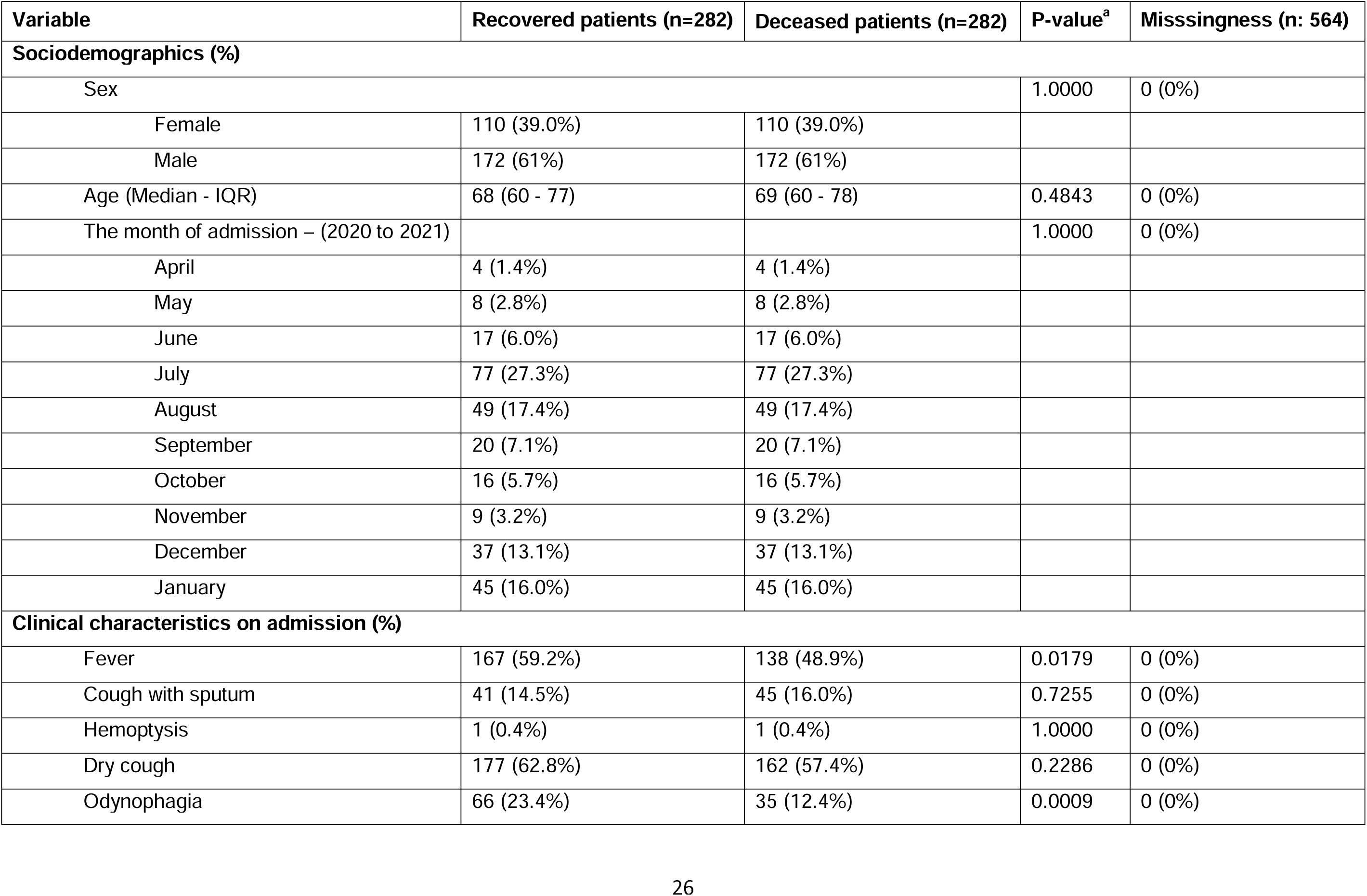
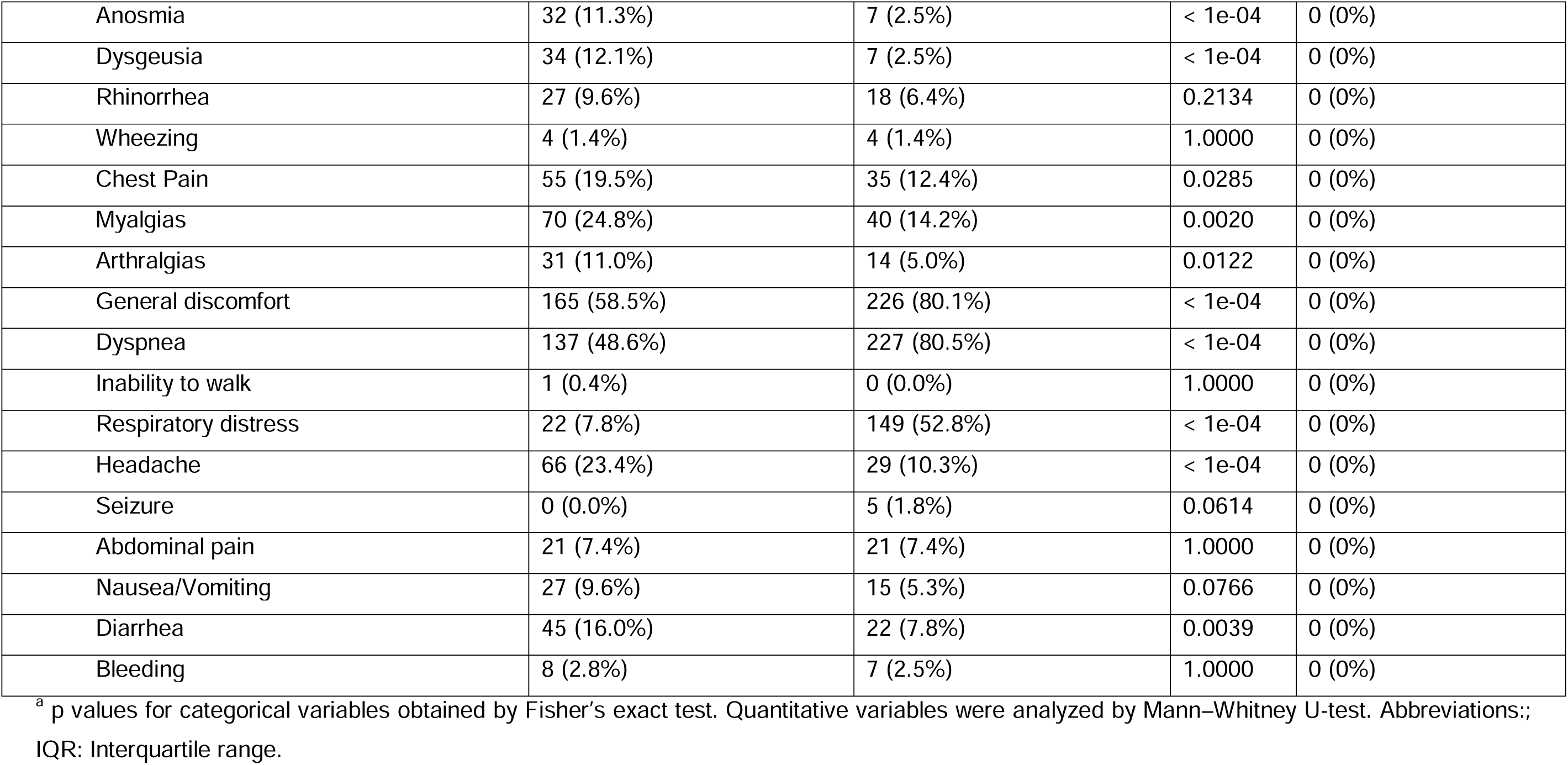
General characteristics of patients diagnosed with COVID-19.

**Table 2.**
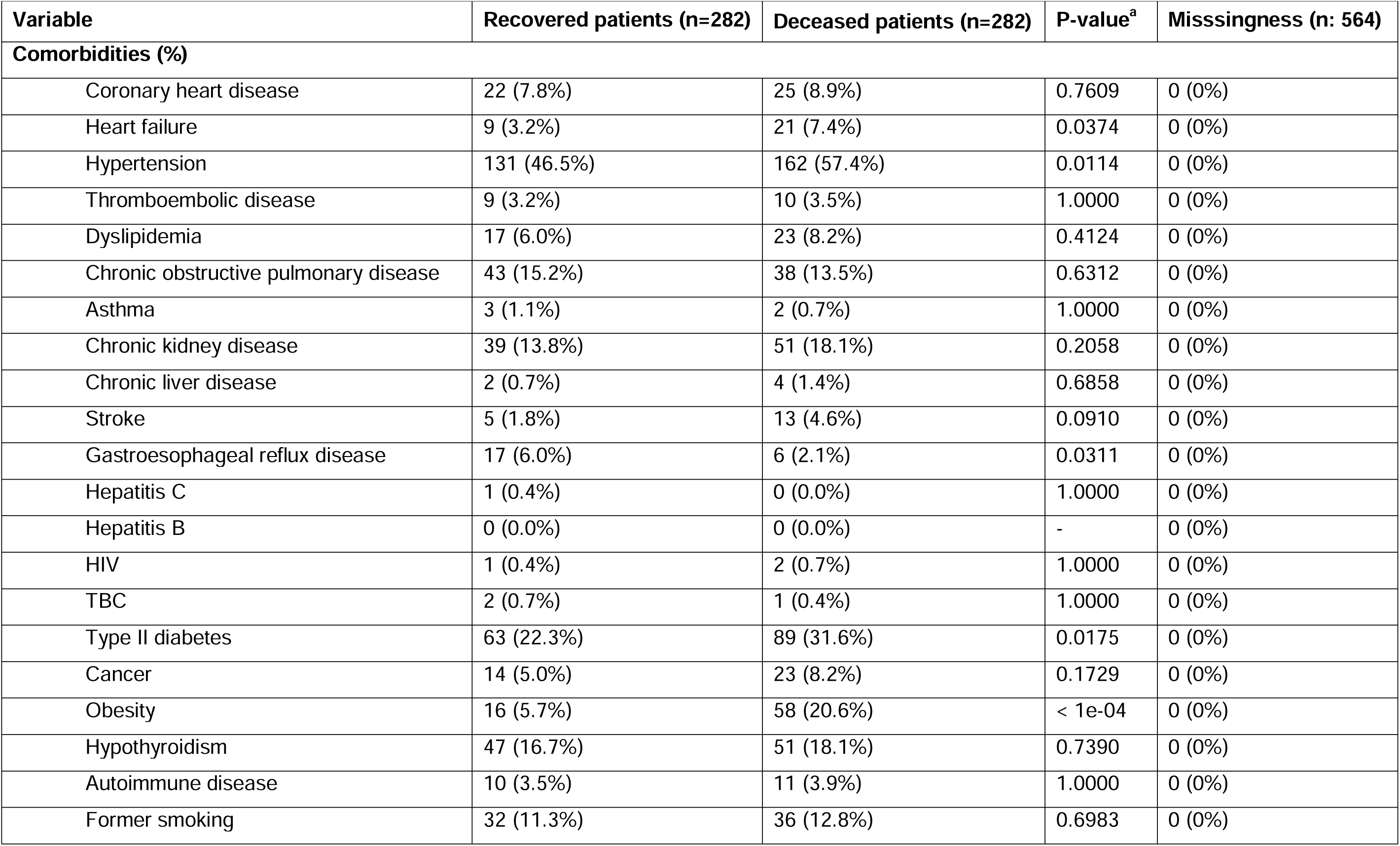
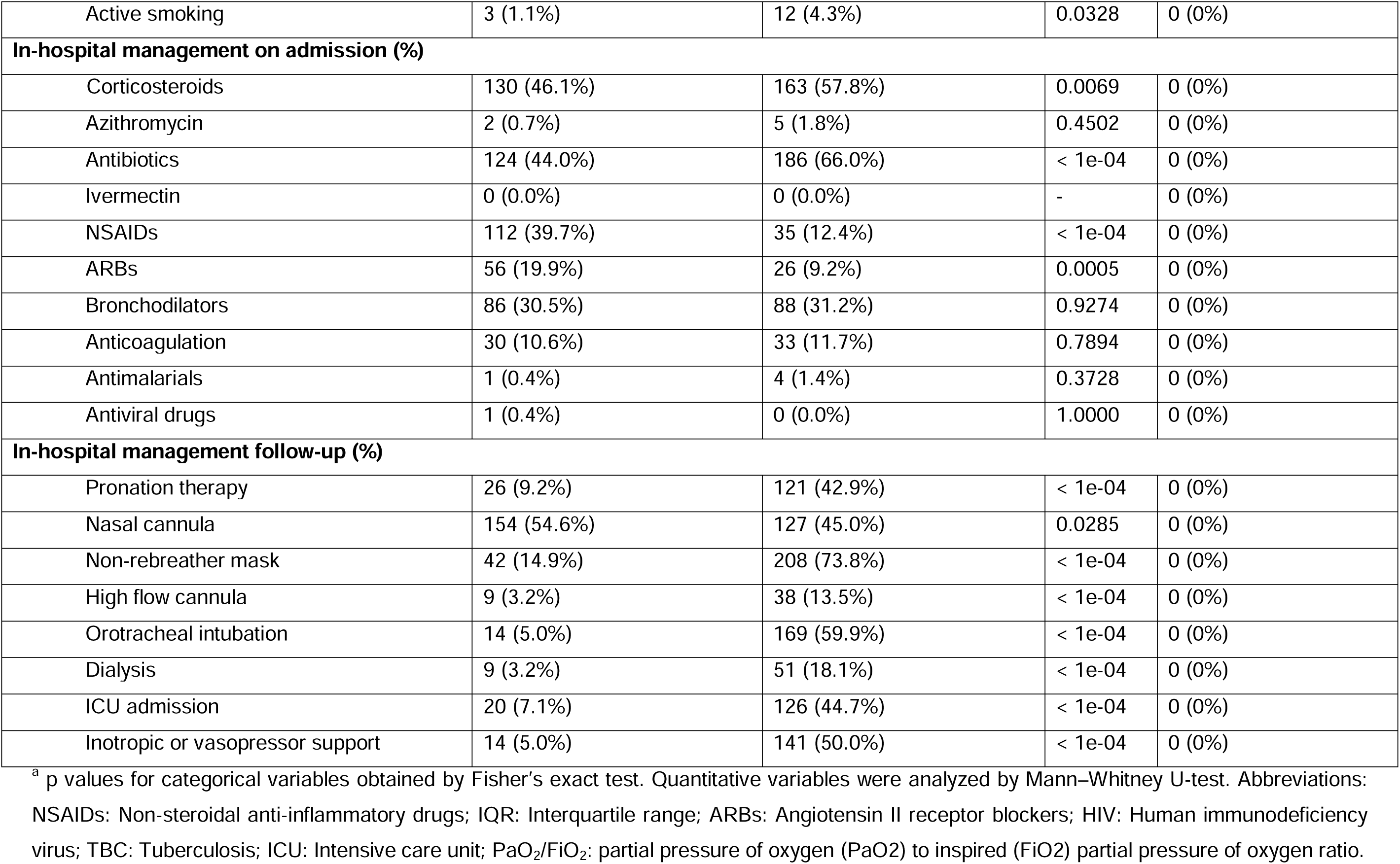
Comorbidities and hospital management of patients diagnosed with COVID-19.

| Variable | Recovered patients (n=282) | Deceased patients (n=282) | P-value<sup>a</sup> | Missingness (n: 564) |
| --- | --- | --- | --- | --- |
| **Comorbidities (%)** |  |  |  |  |
| Coronary heart disease | 22 (7.8%) | 25 (8.9%) | 0.7609 | 0 (0%) |
| Heart failure | 9 (3.2%) | 21 (7.4%) | 0.0374 | 0 (0%) |
| Hypertension | 131 (46.5%) | 162 (57.4%) | 0.0114 | 0 (0%) |
| Thromboembolic disease | 9 (3.2%) | 10 (3.5%) | 1.0000 | 0 (0%) |
| Dyslipidemia | 17 (6.0%) | 23 (8.2%) | 0.4124 | 0 (0%) |
| Chronic obstructive pulmonary disease | 43 (15.2%) | 38 (13.5%) | 0.6312 | 0 (0%) |
| Asthma | 3 (1.1%) | 2 (0.7%) | 1.0000 | 0 (0%) |
| Chronic kidney disease | 39 (13.8%) | 51 (18.1%) | 0.2058 | 0 (0%) |
| Chronic liver disease | 2 (0.7%) | 4 (1.4%) | 0.6858 | 0 (0%) |
| Stroke | 5 (1.8%) | 13 (4.6%) | 0.0910 | 0 (0%) |
| Gastroesophageal reflux disease | 17 (6.0%) | 6 (2.1%) | 0.0311 | 0 (0%) |
| Hepatitis C | 1 (0.4%) | 0 (0.0%) | 1.0000 | 0 (0%) |
| Hepatitis B | 0 (0.0%) | 0 (0.0%) | - | 0 (0%) |
| HIV | 1 (0.4%) | 2 (0.7%) | 1.0000 | 0 (0%) |
| TBC | 2 (0.7%) | 1 (0.4%) | 1.0000 | 0 (0%) |
| Type II diabetes | 63 (22.3%) | 89 (31.6%) | 0.0175 | 0 (0%) |
| Cancer | 14 (5.0%) | 23 (8.2%) | 0.1729 | 0 (0%) |
| Obesity | 16 (5.7%) | 58 (20.6%) | < 1e-04 | 0 (0%) |
| Hypothyroidism | 47 (16.7%) | 51 (18.1%) | 0.7390 | 0 (0%) |
| Autoimmune disease | 10 (3.5%) | 11 (3.9%) | 1.0000 | 0 (0%) |
| Former smoking | 32 (11.3%) | 36 (12.8%) | 0.6983 | 0 (0%) |
| Active smoking | 3 (1.1%) | 12 (4.3%) | 0.0328 | 0 (0%) |
| **In-hospital management on admission (%)** |  |  |  |  |
| Corticosteroids | 130 (46.1%) | 163 (57.8%) | 0.0069 | 0 (0%) |
| Azithromycin | 2 (0.7%) | 5 (1.8%) | 0.4502 | 0 (0%) |
| Antibiotics | 124 (44.0%) | 186 (66.0%) | < 1e-04 | 0 (0%) |
| Ivermectin | 0 (0.0%) | 0 (0.0%) | - | 0 (0%) |
| NSAIDs | 112 (39.7%) | 35 (12.4%) | < 1e-04 | 0 (0%) |
| ARBs | 56 (19.9%) | 26 (9.2%) | 0.0005 | 0 (0%) |
| Bronchodilators | 86 (30.5%) | 88 (31.2%) | 0.9274 | 0 (0%) |
| Anticoagulation | 30 (10.6%) | 33 (11.7%) | 0.7894 | 0 (0%) |
| Antimalarials | 1 (0.4%) | 4 (1.4%) | 0.3728 | 0 (0%) |
| Antiviral drugs | 1 (0.4%) | 0 (0.0%) | 1.0000 | 0 (0%) |
| **In-hospital management follow-up (%)** |  |  |  |  |
| Pronation therapy | 26 (9.2%) | 121 (42.9%) | < 1e-04 | 0 (0%) |
| Nasal cannula | 154 (54.6%) | 127 (45.0%) | 0.0285 | 0 (0%) |
| Non-rebreather mask | 42 (14.9%) | 208 (73.8%) | < 1e-04 | 0 (0%) |
| High flow cannula | 9 (3.2%) | 38 (13.5%) | < 1e-04 | 0 (0%) |
| Orotracheal intubation | 14 (5.0%) | 169 (59.9%) | < 1e-04 | 0 (0%) |
| Dialysis | 9 (3.2%) | 51 (18.1%) | < 1e-04 | 0 (0%) |
| ICU admission | 20 (7.1%) | 126 (44.7%) | < 1e-04 | 0 (0%) |
| Inotropic or vasopressor support | 14 (5.0%) | 141 (50.0%) | < 1e-04 | 0 (0%) |

<sup>a</sup> p values for categorical variables obtained by Fisher's exact test. Quantitative variables were analyzed by Mann-Whitney U-test. Abbreviations: NSAIDs: Non-steroidal anti-inflammatory drugs; IQR: Interquartile range; ARBs: Angiotensin II receptor blockers; HIV: Human immunodeficiency virus; TBC: Tuberculosis; ICU: Intensive care unit; PaO<sub>2</sub>/FiO<sub>2</sub>: partial pressure of oxygen (PaO<sub>2</sub>) to inspired (FiO<sub>2</sub>) partial pressure of oxygen ratio.

**Table 3.** Clinical outcomes and paraclinics on admission of patients diagnosed with COVID-19.

| Variable | Recovered patients (n=282) | Deceased patients (n=282) | P-value<sup>a</sup> | Misssingness (n: 564) |
| --- | --- | --- | --- | --- |
| **Clinical outcomes (%)** |  |  |  |  |
| Renal alterations | 37 (13.1%) | 130 (46.1%) | < 1e-04 | 0 (0%) |
| Co-infection | 20 (7.1%) | 57 (20.2%) | < 1e-04 | 0 (0%) |
| Hematological alterations | 146 (51.8%) | 242 (85.8%) | < 1e-04 | 0 (0%) |
| Thrombotic events | 6 (2.1%) | 18 (6.4%) | 0.0199 | 0 (0%) |
| Neurological alterations | 22 (7.8%) | 31 (11.0%) | 0.2481 | 0 (0%) |
| Cardiac alterations | 18 (6.4%) | 95 (33.7%) | < 1e-04 | 0 (0%) |
| **Paraclinics on admission (Median - IQR)** |  |  |  |  |
| Hemoglobin (g/dL) | 14.8 (13.5-16.2) | 14.4 (12.4-15.8) | 0.0086 | 106 (18.79%) |
| Platelets (Cells/uL) | 206,000 (170,000 - 268,000) | 217,000 (157,000 - 274,000) | 0.8015 | 106 (18.79%) |
| Leukocytes (Cells/uL) | 7,860 (5,620 - 10,285) | 10,670 (6,730 - 14,720) | < 1e-04 | 106 (18.79%) |
| Lymphocytes (Cells/uL) | 1,130 (740 - 1,650) | 860 (585-1200) | < 1e-04 | 106 (18.79%) |
| Neutrophils (Cells/uL) | 5,700 (3,630 - 8,196) | 8,915 (5,267 - 13,112) | < 1e-04 | 107 (18.97%) |
| C-reactive protein (mg/L) | 102.3 (40.2 - 171.2) | 174.7 (85.8 - 267.5) | < 1e-04 | 247 (43.79%) |
| Erythrocyte sedimentation rate (mm/hr) | 22 (22 - 22) | 18 (12 - 20) | 0.3798 | 558 (98.94%) |
| International normalized ratio | 1.06 (1 - 1.17) | 1.07 (0.99 - 1.2) | 0.9196 | 453 (80.32%) |
| Aspartate aminotransferase (U/L) | 37 (27 - 59) | 64.5 (41.8 - 97.5) | 0.0002 | 474 (84.04%) |
| Alanine aminotransferase (U/L) | 30 (21.5 - 54.5) | 46 (30 - 93.3) | 0.0268 | 473 (83.87%) |
| Albumin (gr/dL) | 3.32 (2.97 - 3.48) | 3.21 (2.43 - 3.43) | 0.3918 | 543 (96.28%) |
| Total bilirubin (mg/dL) | 0.64 (0.45 - 0.87) | 0.68 (0.49 - 1.09) | 0.4640 | 422 (74.82%) |
| Blood urea nitrogen (mg/dL) | 18.4 (13.8 - 28.8) | 26.3 (18.4 - 43.2) | < 1e-04 | 166 (29.43%) |
| Creatinine (mg/dL) | 0.99 (0.78 - 1.24) | 1.16 (0.9 - 1.85) | < 1e-04 | 138 (24.47%) |
| Creatine kinase (U/L) | 6.18 (6.18 - 6.18) | 105 (78 - 135) | 0.1432 | 558 (98.94%) |
| D-dimer (mg/dL) | 0.775 (0.41 - 1.74) | 1.52 (0.67 - 4.08) | < 1e-04 | 186 (32.98%) |
| Ferritin (ng/mL) | 878.6 (495.1 - 1,501) | 1,378 (731.2 - 2,475.5) | < 1e-04 | 211 (37.41%) |
| Lactic acid (mmol/L) | 1.44 (1.14 - 1.99) | 2.11 (1.5 - 3.37) | 0.0180 | 443 (78.55%) |
| Lactate dehydrogenase (U/L) | 298 (225.5 - 377.5) | 466 (335.5 - 652) | < 1e-04 | 190 (33.69%) |
| Procalcitonin (ng/mL) | 0.778 (0.55 - 1) | 0.14 (0.09 - 0.19) | 0.0641 | 558 (89.94%) |
| PaO<sub>2</sub>/FiO<sub>2</sub> (mmHg) | 252.6 (211 - 297.80) | 115 (71.5 - 222.15) | < 1e-04 | 155 (27.48%) |
| Oxygen saturation (%) | 92.9 (89.15 - 95.85) | 90.2 (86 - 93,78) | 0.0002 | 131 (23.23%) |

<sup>a</sup> p values for categorical variables obtained by Fisher's exact test. Quantitative variables were analyzed by Mann–Whitney U-test. Abbreviations: NSAIDs: Non-steroidal anti-inflammatory drugs; IQR: Interquartile range; ARBs: Angiotensin II receptor blockers; HIV: Human immunodeficiency virus; TBC: Tuberculosis; ICU: Intensive care unit; PaO<sub>2</sub>/FiO<sub>2</sub>: partial pressure of oxygen (PaO<sub>2</sub>) to inspired (FiO<sub>2</sub>) partial pressure of oxygen ratio.

### Patient and Public Involvement

Patients or the public were not involved in the research’s design, conduct, reporting, or dissemination plans.

### Statistical analyses

Univariate descriptive statistics were performed. Categorical variables were analyzed using frequencies, and continuous quantitative variables were expressed as the median and interquartile range (IQR). The Mann–Whitney U-test or Fisher exact tests were used based on the results. None of the included parameters were subjected to statistical transformation or normalization.

The missing data rates for each variable in our study are shown in Tables 1,2,3 and Figure 1. Most of the missingness was secondary to the lack of standardization at the beginning of the pandemic for laboratory values required in the follow-up and management of the patients. We used multiple imputations by chained equation (MICE) to create and analyze five multiply imputed datasets for variables with less than 80% of data missingness (Table 3). Multiple imputations are considered cutting-edge by methodologists since they enhance accuracy and statistical power when compared to other missing data strategies. Incomplete variables were imputed under wholly conditional specification using the default settings of the MICE 3.14 package (16).

**Fig 1.**
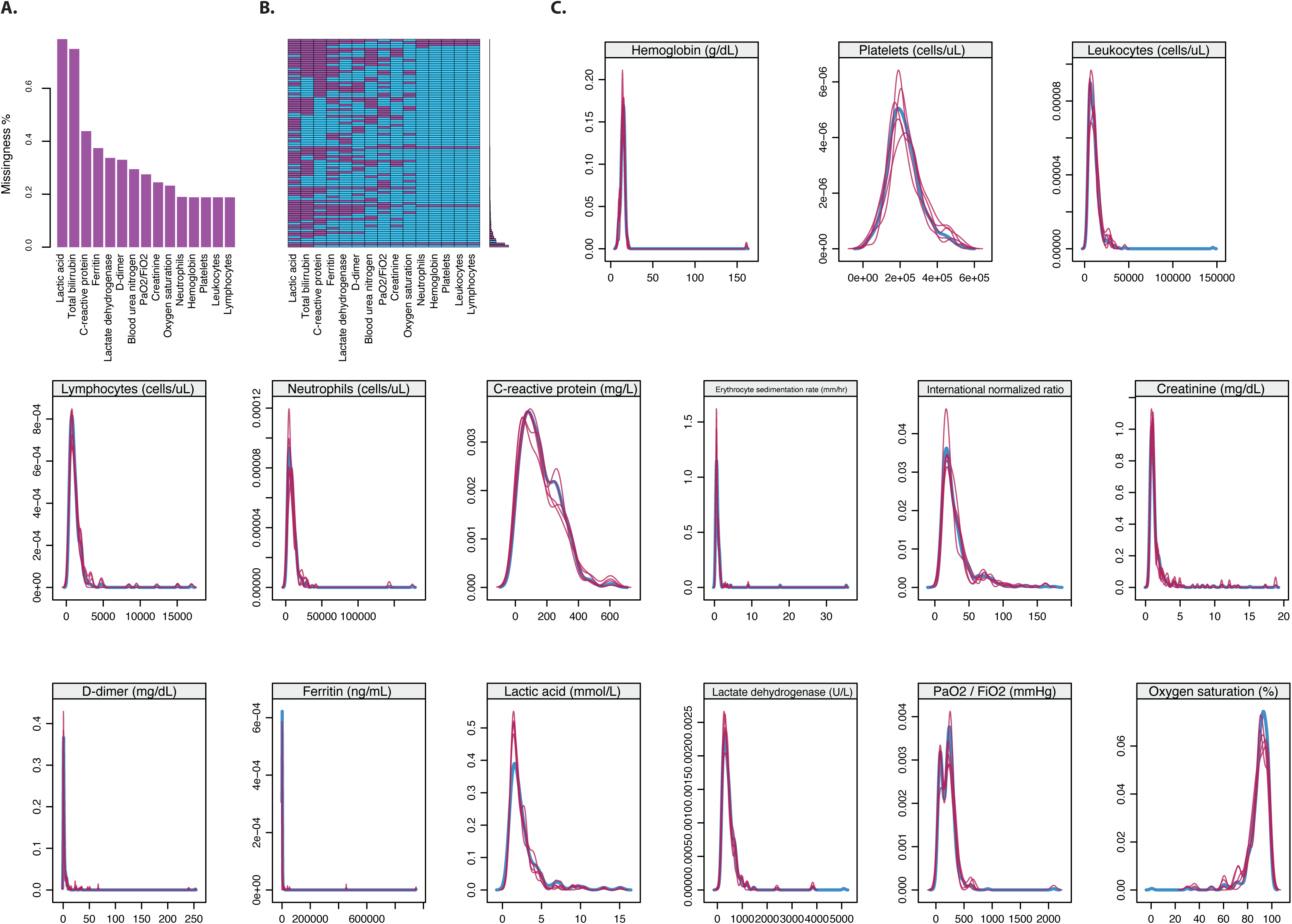
**Missing data and imputation.** **A.** Histogram of frequency of missing variables in the total of patients included (n: 584). **B.** Heatmap for the distribution of missing data. **C.** Distribution of imputed variables by MICE. The red lines correspond to the five imputed datasets, whereas the blue line corresponds to the original dataset. MICE: Multiple imputations by chained equation.

Then, we built classification and regression trees (CART) to evaluate the relationship between clinical variables and mortality in each imputed dataset on admission. This strategy aims to identify, at each partitioning step, the best predictive variable and corresponding splitting value while optimizing a statistical criterion. Variables with a p-value ≤ 0.25 in the bivariate analysis were included in the model. Each model’s confusion matrix was built to determine MICE’s best predictive model from the five imputed datasets and was reported accordingly. The significance level of the study was set to 0.05. Statistical analyses were done using R software version 4.1.2.

## Results

### General characteristics

The general characteristics of patients are shown in Table 1. The median age of the patients and sex were similar between groups. On admission, fever, odynophagia, anosmia, dysgeusia, chest pain, myalgias, arthralgias, headache, and diarrhea were most commonly reported in recovered patients. General discomfort, dyspnea, respiratory distress, and seizures were most common in deceased patients (Table 1). Most of the included patients visited the hospital between July and September 2020.

### Treatment, comorbidities, and outcomes

Heart failure, hypertension, obesity, type II diabetes, and active smoking were most common in deceased patients. On the other hand, gastroesophageal reflux disease (GERD) was mainly reported in recovered patients (Table 1). Deceased patients were more likely to receive corticosteroids and antibiotics upon admission (Table 2). Few patients received antimalarials or antivirals. On admission, deceased patients exhibited paraclinical alterations in inflammatory markers related to hematological, liver, and pulmonary function (Table 3). This was further confirmed during the follow-up, given the higher rates of systemic and organic compromise presented during the hospitalization (Table 3).

### Missing data imputation

The missing values across the variables ranged between 18.79% and 98.94%. The high missingness rate was related to laboratory variables, mainly albumin, creatine kinase, procalcitonin, and the erythrocyte sedimentation rate (Table 3 and Figure 1 A-C), whereas clinical characteristics, in-hospital admission management, and comorbidities did not have missed values. Since the objective of our study was to evaluate the interaction between clinical and paraclinical factors on admission in predicting mortality, we conducted a MICE imputation strategy to include all the cases in multivariate models.

We created and analyzed five multiply imputed datasets for variables with less than 80% of data missingness (Figure 1C). The sensitivity analysis yielded no significant differences between the primary and the five imputed datasets (Table 4). This confirmed that the distribution of imputed data was similar to the original dataset, as well as the stability of the imputation models.

**Table 4.** P-values from sensitivity analysis between the primary dataset and the imputed datasets.

| Variable | Main Vs.<br>Imputed Dataset 1 | Main Vs.<br>Imputed Dataset 2 | Main Vs.<br>Imputed Dataset 3 | Main Vs.<br>Imputed Dataset 4 | Main Vs.<br>Imputed Dataset 5 |
| --- | --- | --- | --- | --- | --- |
| **Paraclinics on admission** |  |  |  |  |  |
| Hemoglobin (g/dL) | 0.4503 | 0.8956 | 0.7301 | 0.8204 | 0.7955 |
| Platelets (Cells/uL) | 0.9705 | 0.7900 | 0.4258 | 0.5854 | 0.8971 |
| Leukocytes (Cells/uL) | 0.9079 | 0.7048 | 0.5637 | 0.4226 | 0.8877 |
| Lymphocytes (Cells/uL) | 0.9808 | 0.7245 | 0.8874 | 0.9945 | 0.8053 |
| Neutrophils (Cells/uL) | 0.8364 | 0.4069 | 0.6002 | 0.2971 | 0.6169 |
| C-reactive protein (mg/L) | 0.7267 | 0.9855 | 0.6387 | 0.8920 | 0.4734 |
| Total bilirubin (mg/dL) | 0.1539 | 0.4270 | 0.7628 | 0.8548 | 0.7584 |
| Blood urea nitrogen (mg/dL) | 0.9875 | 0.5288 | 0.6392 | 0.3416 | 0.9320 |
| Creatinine (mg/dL) | 0.8553 | 0.7842 | 0.9174 | 0.6214 | 0.7608 |
| D-dimer (mg/dL) | 0.9702 | 0.9092 | 0.6611 | 0.9079 | 0.7723 |
| Ferritin (ng/mL) | 0.7442 | 0.5230 | 0.1959 | 0.8348 | 0.4637 |
| Lactic acid (mmol/L) | 0.2466 | 0.2657 | 0.3806 | 0.4587 | 0.2240 |
| Lactate dehydrogenase (U/L) | 0.7523 | 0.4675 | 0.9154 | 0.7937 | 0.5737 |
| PaO<sub>2</sub>/FiO<sub>2</sub> (mmHg) | 0.9339 | 0.9456 | 0.6239 | 0.7643 | 0.5535 |
| Oxygen saturation (%) | 0.6625 | 0.9327 | 0.4655 | 0.6727 | 0.8556 |

<sup>a</sup> p values for quantitative variables were analyzed by Mann–Whitney U-test. PaO<sub>2</sub>/FiO<sub>2</sub>: partial pressure of oxygen (PaO<sub>2</sub>) to inspired (FiO<sub>2</sub>) partial pressure of oxygen ratio.

### A multivariate predictive model for mortality

After imputation, we aimed to evaluate the interaction of multiple variables in predicting mortality on admission. We estimated CART models using the variables with p-values ≤ 0.25 from the bivariate analysis for each imputed dataset. We constructed a confusion matrix for each CART model to estimate the best-fitted model to the data. After this analysis, we selected the best model based on estimated accuracy (Figure 2).

**Fig 2.**
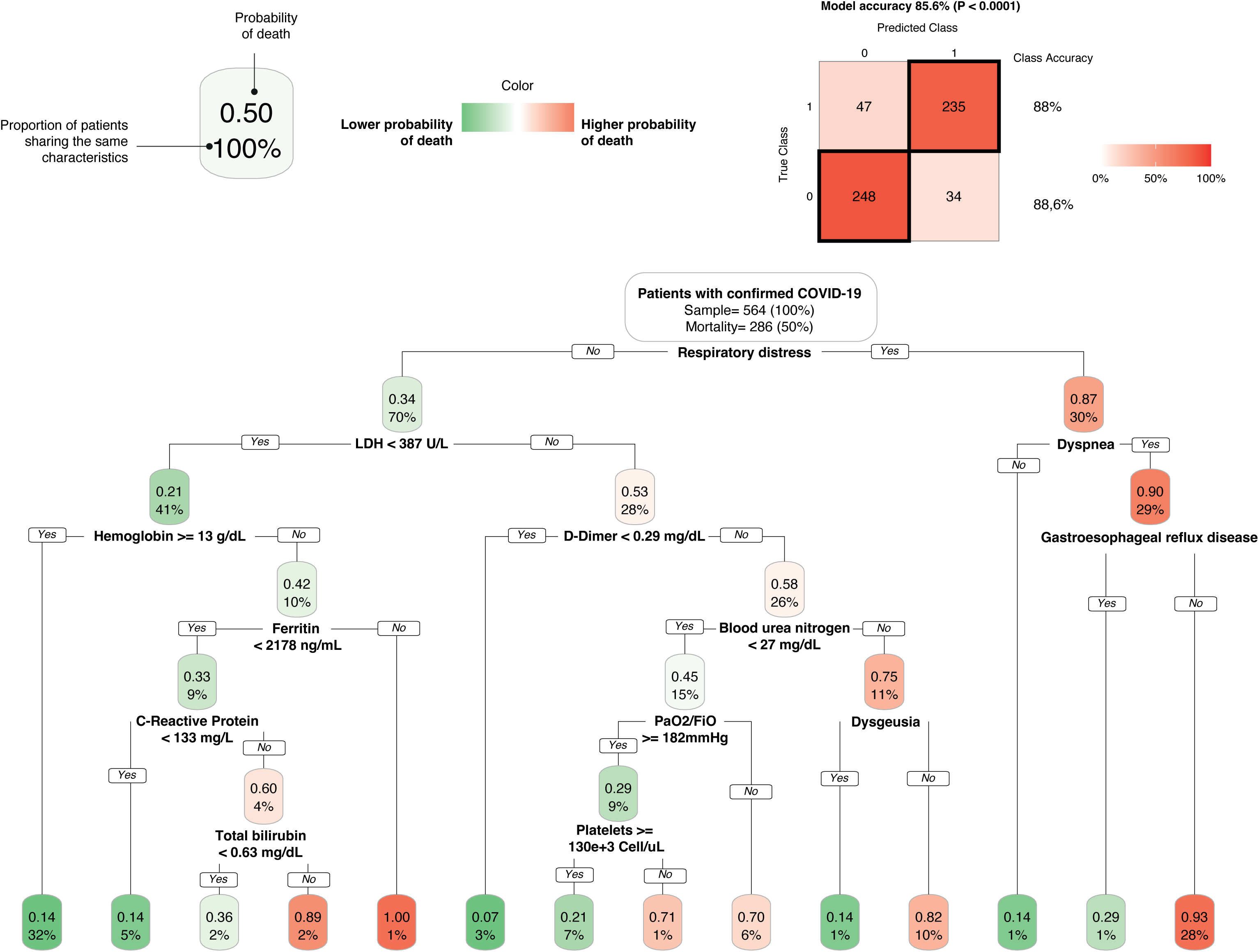
**Classification and regression trees (CART).** This strategy estimated a predictive model and 14 clinical profiles, including respiratory distress, LDH, dyspnea, hemoglobin, D-dimer, ferritin, blood urea nitrogen, C-reactive protein, PaO_2_/FiO_2_, dysgeusia, total bilirubin, platelets, and gastroesophageal reflux disease. LDH: lactate dehydrogenase; PaO_2_/FiO_2_: partial pressure of oxygen (PaO_2_) to inspired (FiO_2_) partial pressure of oxygen ratio.

The analysis revealed that multiple variables interacted in the prediction of mortality. Respiratory distress on admission was the first splitting variable from the tree. Then, the second node was determined by LDH and dyspnea. The former interacted with hemoglobin, D-dimer, ferritin, blood urea nitrogen (BUN), C-reactive protein (CRP), partial pressure of oxygen (PaO_2_) to inspired (FiO_2_) partial pressure of oxygen ratio (PaO_2_/FiO_2_), dysgeusia, total bilirubin levels (TBIL), and platelets. The latter interacted with GERD (Figure 2).

## Discussion

A few published papers have previously characterized the risk factors associated with COVID-19 mortality in Latin American patients (17,18) Therefore, the present study highlights multiple variables, including laboratory abnormalities and clinical features associated with COVID-19 mortality. The main findings in our study were significant associations between TBIL, ferritin, D-Dimer levels, dyspnea, GERD, and increased risk of mortality in COVID-19 patients. In contrast, dysgeusia was associated with a better prognosis. Other trends were found between BUN, respiratory distress, platelet count, PaO2/FiO2, and CRP.

Increased TBIL was associated with poor COVID-19 outcomes. This is consistent with previous findings (19) which suggest that high TBIL may reflect a severe level of hepatic injury among severely ill COVID-19 patients, possibly due to direct cytopathic effect, immune-mediated effects, hypoxia-induced changes, microvascular thrombosis, among others (20,21). It is not surprising that some authors have also considered that TIBL at admission is directly correlated with the hospital progression of COVID-19 (20).

Serum ferritin has been cited as one of the mortality indicators in COVID-19 patients due to its ability to assess intracellular iron status (22,23). These events are common in the pathogenesis of uncontrolled inflammation and massive cytokine release, supporting the hypothesis that hyper-inflammation is a possible pathogenic mechanism in COVID-19 and, therefore, the rise of serum ferritin level findings (23). Multiple studies have also suggested that higher levels of CRP on admission are linked to disease progression, severity, and death (24–26). However, we found that CRP interacts with other variables related to inflammation and organ damage implicated in mortality.

The D-dimer antigen is a unique marker of fibrin degradation that may indicate infection-related coagulation effects (27,28). Furthermore, previous studies have found that critically ill patients with COVID-19 have extremely high D-dimer levels, which can lead to clotting disorders and peripheral microthrombi formation (29). In this study, we found that higher levels of D-dimer at admission were associated with increased mortality risk in COVID-19 patients. This is consistent with other studies that have found D-dimer as another crucial prognostic factor in estimating mortality in COVID-19 patients (30).

Similar results from other studies also elicited dyspnea as a significant clinical variable for mortality prediction among COVID-19 patients (31). A meta-analysis study by He et al. showed that dyspnea was the main difference between mild and severe COVID-19 [40], and another study confirmed this observation (32). On the other hand, in patients with gastrointestinal symptoms during COVID-19, GERD appeared to be a protective factor for acute respiratory distress syndrome (ARDS), and mortality possible due to a conversely increased acidic environment associated with GERD that suppresses COVID-19 viral load at the gastrointestinal point of entry, favoring a milder disease course (33). In our study, similar results were obtained; COVID-19 patients without GERD had an increased mortality risk. Interestingly, dysgeusia was associated with lower odds of death in our study. This might be related to a different inflammatory profile with a better local immune response, which could limit the spread of the virus in the body, resulting in less severe disease and a strong local inflammatory response that could mainly affect taste receptors. However, there is still a lack of information regarding dysgeusia, perhaps because of the heterogeneity in how it has been assessed and defined (34). On the other hand, previous studies have focused on anosmia as a protective factor for mortality (35,36).

A significant association was found between BUN and mortality risk among COVID-19 patients. Other studies also supported our results since BUN has been previously considered a death-related feature (37). Although COVID-19 impacts mainly the lung, it can also affect the kidney, and the increase in BUN may reflect kidney injury along with other biomarkers. Moreover, kidney involvement in severe COVID-19 patients has been frequently observed (37,38).

The glycolytic enzyme LDH has long been identified as an inflammation biomarker that plays a crucial role in the anaerobic glycolysis pathway and increases in the bloodstream under conditions of membrane instability (39). In most studies, authors have concluded that LDH is a deleterious prognostic biomarker with high accuracy for predicting in-hospital mortality in severe and critically ill patients with COVID-19 (40,41). In addition, thrombocytopenia has also been reported in COVID-19 patients and is considered a potential risk factor for mortality in this group of patients (42,43). The PaO2/FiO2 partial pressure of oxygen has also been widely used to diagnose and assess the severity of patients with ARDS (44,45). Since several factors have been related to the incidence of mortality in COVID-19, the interaction among them may provide better insights into predicting deleterious outcomes. A similar study in Bogota found that older age, low S/F, and high LDH on admission were predictors of mortality (13). This study assessed the interaction of variables by CART analysis, yielding five plausible groups. However, it is unclear whether the multivariate model included all the patients since information on missing data is unavailable, and the model accuracy is unknown.

In contrast, Afrash et al.(46) developed hybrid machine-learning algorithms to predict mortality. Authors found that the mixture of variables such as length of stay (LOS), age, cough, respiratory intubation, dyspnea, cardiovascular disease, leukocytosis, BUN, CRP, and pleural effusion yielded a high accuracy (90%), specificity (83%), and sensitivity (97%). However, it includes variables that depend on the patient follow-up (i.e., LOS), thus hindering its applicability during admission to the emergency room, that is, during the early stages of the disease.

Several manuscripts have developed similar approaches involving different variables in the prediction of mortality; some of them include dyspnea (46,47), BUN, platelet count (47,48), sex (47), age (46,48–51), cough (46), weight (49), cardiovascular disease (46), orotracheal intubation (46), and pleural effusion (46,49), respiratory rate (47), fraction of inspired oxygen (47), blood oxygen saturation (47,48) pH (47), aspartate aminotransferase levels (47), estimated glomerular filtration rate (47), lymphocyte count (49,52), white blood cell count (46,48), creatine (51) lactic acid (52) and serum calcium (52). However, most of them do not focus on the interaction of such variables on hospital admission, and their interpretability by clinicians is difficult. In this line, our study provides a novel, biologically plausible, and reliable model with clinical applicability in the emergency room during patient admission. Further studies are necessary to evaluate whether our model may help to predict the efficacy of therapeutic strategies or outpatient mortality (during post-COVID syndrome).

Study limitations must be acknowledged. It was a single-center retrospective study based on clinical records. This could have prone our study to reporting bias, for example, on time from symptoms onset to consultation or clinical features. In addition, the therapeutic strategies changed during the pandemic; however, our study was matched by month of consultation, making this variable an unlikely confounding/interaction factor for our results. The objective of our study was to uncover other risk factors for mortality besides sex and age. In this line, the adjustment for age, sex, and month of consultation allowed the discovery of new interactions for mortality prediction. The lack of follow-up beyond the hospitalization in recovered patients could have offered new insights into mortality beyond the clinical settings. Another potential shortcoming of the present study is that latent autoimmunity, especially the positivity for anti-IFN-α antibodies, was not evaluated. Further analysis involving such variables could improve our model’s reliability and predictive accuracy.

## Conclusions

Our study demonstrates the highly complex interactions among different risk factors to predict mortality in COVID-19, in addition to age and sex. This predictive approach may also provide new insights into the tailored management of this illness in several clinical settings, specifically in Colombian clinical settings. This study should encourage the follow-up of recovered patients beyond hospitalization and despite their clinical status during acute COVID-19.

## Ethics approval and consent to participate

The institutional review board of Clínica de Occidente approved the study design. Written informed consent was not obtained from participants since the study was based on a retrospective review of clinical records. It was a low-risk study according to the resolution 8430 of 1993 from the Ministry of Health of Colombia. The study was conducted following the principles stated in the Declaration of Helsinki and Good Clinical Practice guidelines.

## Consent to publish

None of the patients were required to consent to publish, given the study’s retrospective nature.

## Author Contributions Statement

Conceptualization: JMA; Acquisition of data: LCD, MR, YR, MH, SB, PSG, NAC, ME, SG, VB, VM, JEC, EZ; Methodology: JMA, LCD, MR; Statistical Analysis: MR; Funding acquisition: JMA, CRS; Center Coordination: YR, MR; Writing & editing: LCD, MR, CRS, YAA, DMM. All authors read and approved the final version of this manuscript.

## Data Availability

All data produced in the present study are available upon reasonable request to the authors

## List of Abbreviations

ARBs: Angiotensin II receptor blockers.
ARDS: Acute respiratory distress syndrome.
BUN: Blood urea nitrogen.
CART: Classification and regression trees.
COVID-19: Coronavirus disease 2019.
CRP: C-reactive protein.
FiO_2_: Fraction of inspired oxygen.
GERD: Gastroesophageal reflux disease
HIV: Human immunodeficiency.
ICU: Intensive care unit.
IL-1: Interleukin 1.
IL-6: Interleukin 6.
IQR: Interquartile range.
LDH: Lactate dehydrogenase.
LOS: Length of stay.
Masl: Meters above sea level.
MICE: Multiple imputations by chained equation.
MV: Mechanical ventilation.
NSAIDs: Non-steroidal anti-inflammatory drugs.
PaO_2_/FiO_2_: partial pressure of oxygen (PaO_2_) to inspired (FiO_2_) partial pressure of oxygen ratio.
RT-PCR: Reverse transcriptase-polymerase chain reaction.
S/F: Ratio of oxygen saturation to fraction-inspired of oxygen.
SARS-CoV-2: Severe acute respiratory syndrome coronavirus 2.
TBC: Tuberculosis.
TBIL: Total bilirubin levels.
TNFα: Tumor necrosis factor-α.
WHO: World health organization.

## Acknowledgments

The authors would like to thank the medical staff of Clinical del Occidente, Dr. Ivan Torres, and Maria Claudia Fuentes, for their assistance at the beginning of the study.

## Availability of data and materials

Data will be available upon request to the corresponding author.

## About the Author

Dr. Camacho-Domínguez is a research assiant in autoimmune diseases at Center for Autoimmune Diseases Research (CREA), School of Medicine and Health Sciences, Universidad del Rosario, Bogota, Colombia. Her primary research interests include infectious diseases, immunology, and rheumatology.

